# Incidence, risk factors and outcomes for neonatal sepsis in The Gambia: Descriptive cohort analysis embedded in a randomised clinical trial

**DOI:** 10.1101/2025.10.04.25336976

**Authors:** Helen Brotherton, Usman N Nakakana, Bully Camara, Nathalie Beloum, Joquina Chiquita Jones, Fatoumata Sillah, Shashu Graves, Madikoi Danso, Saffiatou Darboe, Solomon Umukoro, Abdoulie Bojang, Ellen Sambou, Isatou Jagne, Buntung Ceesay, Momodou Barry, Kady Kassibo, Aminata Cham, Yusupha Njie, Ebrahim Ndure, Kebba Manneh, Christian Bottomley, Halidou Tinto, Umberto d’Alessandro, Anna Roca

## Abstract

**Background:** Neonatal sepsis is a major contributor to adverse neonatal outcomes in West Africa. High quality data on risk factors for sepsis in this region is limited, yet important for surveillance, prevention and management of newborns at greatest risk. This study aimed to describe the clinical epidemiology of sepsis for health-facility born neonates in an urban West African setting.

**Methods:** This study comprised secondary data analyses from the Gambian cohort of the PregnAnZI-2 randomised clinical trial. Relatively healthy pregnant women and their offspring who delivered at two urban health facilities were included, with a combination of active and passive surveillance until 28 days. Neonatal sepsis was defined as suspected if clinical and laboratory (abnormal FBC or CRP) criteria were met, and confirmed if blood-culture was positive. A novel conceptual framework informed logistic regression models to identify 1) factors associated with neonatal sepsis and 2) contribution of sepsis towards neonatal mortality.

**Results:** A total of 6515 neonates were included. The health-facility based incidence of neonatal sepsis was 20.2 cases/1000 live births (N=131 cases), predominantly early-onset (<3 days)(15.7 cases/1000 livebirths). Confirmed sepsis accounted for 22% (29/131) of all cases, with *Burkholderia cepacia* and *Staphylococcus aureus* the most prevalent bacteria in 24% (7/29) of confirmed sepsis each. Risk factors for sepsis included low 1-minute Apgar score (aOR 13.2, 95% CI 8.40-20.73), pre-labour maternal fever (aOR 5.0, 95% CI 1.15-21.69), easily recognisable congenital malformation (aOR 3.39, 95% CI 1.55-7.38) and low-birth weight (aOR 2.85, 95% CI 1.75-4.65). 40.7% of all neonatal deaths in the cohort occurred in neonates with sepsis, with 40-fold increase in mortality compared to neonates without sepsis (OR: 39.98, 95% CI 22.5 – 71.1).

**Conclusion:** Sepsis, especially early onset, is a major morbidity for health facility born neonates delivered following relatively healthy pregnancy in urban Gambia, with high associated mortality. We identify neonatal phenotypes (low birth weight, newborns with low 1-minute apgar scores, or those with easily recognisable congenital malformations) who may benefit from enhanced postnatal surveillance or antibiotics to prevent or treat neonatal sepsis and reduce neonatal mortality.

**Trial Registration:** NCT03199547: Clinicaltrials.gov. Registered on 23^rd^ June 2017

## BACKGROUND

An estimated 3 million neonates develop sepsis annually[1], predominantly in low-and middle-income countries (LMIC). The global incidence of neonatal sepsis is increasing, likely due to recent estimates including more studies from LMICs and a lack of recent data from high-income country (HIC) settings[2]. Severe infections, including sepsis, are the third most common cause of neonatal deaths globally and the leading cause of death amongst newborns aged 2-7 days old[3]. Africa has the highest incidence of neonatal sepsis of all WHO regions, estimated at 5243 cases/100,000 live births annually[2], although precise within-country estimates are currently lacking. Neonatal sepsis has a substantial negative impact on child survival, health and neurodevelopment. Infections, including sepsis, account for nearly one-quarter of neonatal deaths in Africa[4], with long-term sequelae in survivors resulting in loss of 5.3 – 8.7 million DALYs annually with an associated economic cost of > $10 billion / year[5].

Accurate clinical diagnosis of neonatal sepsis is challenging in all settings due to non-specific features, but especially in low-resource health facilities, where access to diagnostics for raised inflammatory markers and microbiological culture are frequently limited. Hence, recognition of at-risk maternal and neonatal phenotypes is often used to guide clinical decision-making for antibiotic prophylaxis and treatment. Risk factors for sepsis are well described in HICs, with established algorithms for identification and management of newborns at greatest risk (e.g., maternal fever, prolonged rupture of membranes, previous group B streptococcus invasive disease)[6]. However, there is a lack of context-specific understanding of risk factors in low-resource West African settings, where differences exist in the aetiology of neonatal bacterial infections[7], epidemiology of maternal bacterial carriage[8], environmental exposures and health systems context.

Understanding context-specific risk factors is foundational for developing clinical algorithms and guidelines to prevent, detect and treat at-risk newborns in regions with greatest sepsis incidence, such as West Africa. A recent systematic review and meta-analysis of African studies (32 studies, 14 from West Africa, n=22,731) identified 15 maternal and neonatal risk factors, some not previously included in WHO guidelines [9]. Despite the large population studied, this review highlighted the high risk of bias, poor quality of most studies and heterogeneity in study design and sepsis definitions, signalling the need for higher quality research in this field[9]. The NeoTree project in Zimbabwe recently developed a neonatal sepsis predictive model which included several maternal and neonatal risk factors identified within an East African population of term neonates[10]. However, the model offers only a limited capacity to detect early-onset sepsis, as indicated by an area under the ROC curve of 0.74. Additionally, the specificity of the NeoTree model is only 11% at a risk score cut-off point that achieves 95% sensitivity, limiting ability to target treatment and risking over-use of antibiotics, driving anti-microbial resistance. Hence, more specific, evidence-based risk-factor algorithms are needed for the highest burden settings.

This study aimed to investigate the clinical epidemiology of neonatal sepsis among a cohort of newborns delivered at two urban health facilities in The Gambia, West Africa. Specific objectives related to describing the incidence, risk factors, clinical presentation, aetiology and contribution of sepsis towards neonatal mortality.

## Methods

### Study design

This cohort study comprised secondary data analyses from the PregnAnZI-2 trial (clinicaltrials.gov ref: NCT03199547), a phase III, double-blind, placebo-controlled randomised clinical trial conducted in The Gambia and Burkina Faso from 2017 to 2021. Azithromycin or placebo were administered to women in labour to assess effect on a composite primary outcome of neonatal mortality or sepsis [11]. The PregnAnZI-2 trial demonstrated a protective effect of intra-partum azithromycin on maternal mastitis and puerperal fever in addition to clinical and bacteriological confirmed neonatal skin infections and need for antibiotics, but no effect on incidence of neonatal sepsis or mortality [12]. Hence, this study used data from both the intervention and control trial groups. The aetiology of blood-culture confirmed neonatal sepsis showed slight but non-significant between-group variation, with more gram-negative bacteria in the azithromycin group (0.15%) versus the placebo group (0.22%)(risk difference −0.07, 95% CI-0.23 to 0.10)[12]. Only the Gambian cohort were included in these post-hoc analyses as most neonatal sepsis cases occurred in The Gambia (131/157, 83%), although rates of neonatal mortality were similar between countries, suggesting that sepsis episodes in Burkina Faso may have been missed [12].

### Setting

Participants were recruited between 2017 and 2021 at two urban government health facilities in Western Gambia: Serekunda Health Centre and Bundung Maternal and Child Heath Hospital (BMCHH) [11]. Serekunda Health Centre provided primary level health services with no facility for emergency obstetric or specialist neonatal inpatient services. Hence, unwell mothers and newborns were referred to Kanifing General Hospital (3.4Km distance away) or Edward Francis Small Teaching Hospital (EFSTH, 15.5Km distance away), where WHO level 2+ small and sick newborn care was available [13]. BMCHH had facilities for emergency obstetric management and a neonatal unit providing WHO level 2 newborn care[13] including oxygen, IV fluids and antibiotics. The Clinical Services Department at MRC Unit The Gambia at LSHTM (MRCG) was also a referral site for neonates requiring admission from the community and provided level 2 newborn care. The neonatal mortality rate at population level in The Gambia during the study period ranged from 28/1000 livebirths (2017)[14] to 25/1000 livebirths (2021)[4].

### Participants

This study included pregnant women aged ≥16 years and their live-born newborns who were recruited to the PregnAnZI-2 clinical trial in The Gambia. Maternal exclusion criteria included: Known HIV infection; Acute or chronic condition at randomisation such as prolonged rupture of membranes, maternal fever or diabetes mellitus; Planned caesarean section or anticipated referral to tertiary health facility; Known severe congenital malformation; macrolide allergy or use of medication known to prolong QT interval in preceding two weeks [11]. PregnAnZI-2 neonatal participants who were admitted to hospital for reasons other than sepsis were entirely excluded from this post-hoc study. This was done in recognition of the low specificity of neonatal sepsis diagnosis using clinical features and inflammatory tests and to avoid over classification of neonates with sepsis.

### Study procedures, data collection and follow-up

Sensitisation and informed consent procedures took place antenatally with confirmation of informed consent, screening and recruitment during labour. Socio-demographic and clinical data were collected by trained personnel during labour and the postpartum hospital admission (typically 6 – 24 hours after delivery) using medical records, direct observations and maternal interviews. Birth weight was measured using a digital, calibrated scale. All newborns were examined by a trained research clinician prior to hospital discharge, with documentation of vital signs. Standard newborn care at recruitment sites prior to discharge included umbilical cord care (surgical spirit), eye care (tetracycline), initiation of breast feeding, thermal care and measurement of vital signs (heart rate, temperature, respiratory rate), with discharge only if vital signs were normal. Participants received BCG, oral polio and Hepatitis B vaccinations according to the national Expanded Programme on Immunisation schedule.

Active follow-up occurred at postnatal day 28 via a home visit. Passive case detection of adverse events, including sepsis, took place from hospital discharge to 28 postnatal days and consisted of participants making unscheduled visits to the trial sites or contacting the trial team with concerns about their newborn. Mothers were provided with information about signs of sepsis before discharge along with research team contact numbers and mobile phone credit to encourage communication. Trained research clinicians assessed newborns for clinical sepsis and advised admission, referral to another facility or outpatient treatment according to clinical judgement, with ongoing follow-up and data collection by the trial team [11]. Hospitalised neonates with sepsis suspected on clinical grounds by the research clinician or paediatrician underwent venepuncture for blood culture, Full Blood Count (FBC) and C-Reactive protein (CRP) by trained study personnel according to study specific procedures and prior to administration of antibiotics where possible. Blood cultures were obtained using aseptic technique with minimum 1ml blood volume, using *BACTEC Peds* Plus™/F vials. Blood was collected into Ethylene Diamine Tetraacetic Acid (EDTA) and heparinised sample bottles for measurement of FBC and CRP respectively. Clinical management of neonatal sepsis was as per hospital guidelines and at the discretion of the facility medical staff, with ampicillin and gentamicin for early onset sepsis and either cefotaxime, flucloxacillin or ampiclox (ampicillin - cloxacillin) and gentamicin for late-onset sepsis, depending on clinical presentation.

### Laboratory procedures

All samples were transported immediately to MRCG clinical laboratories (ISO 15189 accredited and GCLP certified). Blood cultures were prospectively processed using the BACTEC 9050 BD machine. Samples exhibiting positive signal underwent Gram-staining and sub-culture as per standard culture methods with characterisation using appropriate biochemical reagents and species identification using API20E (Enterobacterales), API20NE (other non-enteric Gram-negative bacteria) or Staphaurex Plus (*Staphylococcus aureus*)[11]. Antimicrobial susceptibility patterns were determined by Kirby-Bauer disk diffusion on Mueller-Hinton agar and zone sizes interpreted according to the relevant Clinical Laboratory Standard Institute guidelines on antimicrobial agents. Antibiotic susceptibility data will be reported separately with consideration of how the PregnAnZI-2 trial antimicrobial intervention may have influenced antimicrobial resistance patterns. FBC (white cell count, neutrophil count and platelets) results were generated using either the CELL DYN Ruby 5-part differential analyser or the 3-part Medonic M-Series Haematology analyser. The VITROS 350 Biochemistry analyser, COBAS C111 or Randox Daytona Plus Chemistry automated analyser were used to generate qualitative CRP levels (>10 mg/L), with validation checks showing comparable results between systems.

### Outcome measures and definitions

The primary endpoint was neonatal sepsis, both suspected and confirmed cases, as defined below. Neonatal sepsis was further classified as early onset sepsis (<3 days) and late onset sepsis (3 to 28 days) and considered as secondary endpoints for incidence calculations.

Definitions of suspected neonatal sepsis were consistent with the PregnAnZI-2 trial [11, 12] and other trials of neonatal infection interventions conducted in Africa [15]. Early onset suspected sepsis was defined as onset of symptoms at less than 3 days of age (date and time of admission taken as proxy) with either respiratory distress or ≥2 other clinical criteria and ≥1 laboratory criteria (Figure 1). Late onset suspected sepsis was defined as onset between 3 and 28 days of age with ≥1 laboratory criterion plus either a) 2 features of respiratory distress; or b) 1 criterion of respiratory distress and another clinical feature; or c) ≥2 non-respiratory clinical criterion (Figure 1). Confirmed sepsis was defined as hospitalised neonates with isolation of a recognised neonatal pathogen from a blood culture, regardless of whether criteria for suspected sepsis were met. Bacterial blood culture contaminants were pre-defined as Coagulase negative *Staphylococcus, Micrococci or Bacillus* spp., Diphtheroids, *Viridans streptococci, Anthrobacter* spp., *Rhodococcus* spp. [11, 12].

**Figure 1.**
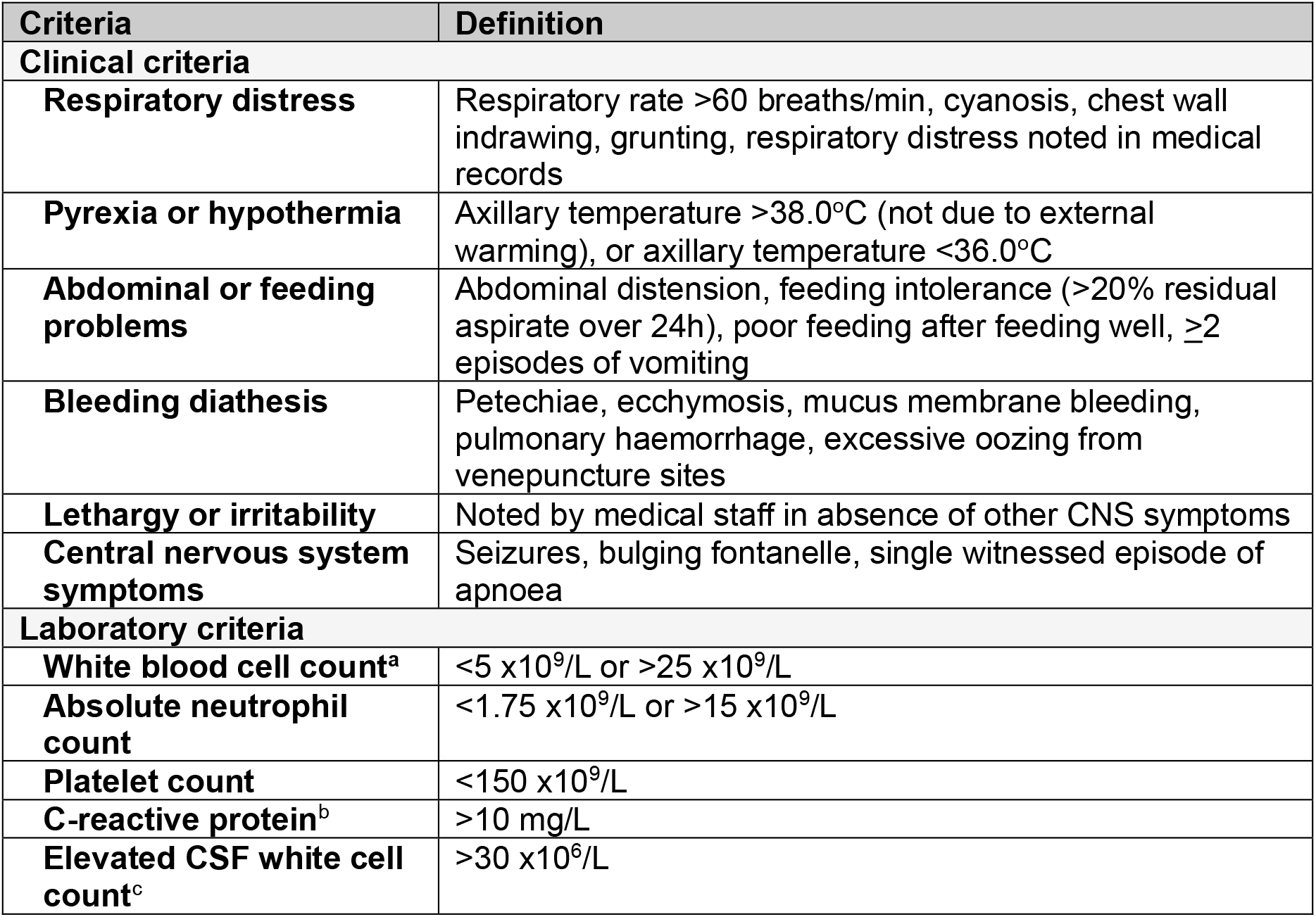
Clinical and laboratory criteria for diagnosis of suspected neonatal sepsis[11]. Abbreviations: CNS = Central Nervous System; CSF = Cerebral-spinal fluid a) In absence of receiving corticosteroids; b) CRP available for early onset sepsis only; c) In absence of significant CSF red cells

### Data management and statistical analysis

All data were collected using electronic data capture on REDCap software with consistency checks, data validation and cleaning carried out as per PregnAnZI-2 trial procedures. Data were pseudo-anonymised with each participant assigned a unique identifier.

All analyses were conducted with Stata version 17.0 (Statacorp). The incidence of neonatal sepsis was determined as the proportion of neonates meeting the study definition of suspected or confirmed sepsis (cases) out of the total number of live-born neonates in our cohort (excluding neonates admitted for another reason). In addition, we also determined the incidence rates of early- and late-onset sepsis for all sepsis cases and confirmed sepsis cases. An exploratory analysis of clinical and laboratory features of neonates with sepsis depending on blood culture status was conducted. This was done to understand the clinical phenotype of septic neonates, in recognition of the non-specific signs of sepsis and overlap with other conditions, especially intrapartum related asphyxia (IRA). Clinical features were compared between those with blood culture obtained (whether the blood culture was positive, negative, or contaminated) versus no blood culture, using Fishers exact test for categorical variables and Student t test for continuous variables.

A comprehensive novel conceptual framework was developed, based on biological plausibility and existing evidence for risk factors and pathophysiology of neonatal sepsis in Africa (Figure 2). The framework considers both protective and adverse factors that may contribute towards neonatal sepsis, covering distinct phases (antenatal, intrapartum and postnatal) as factors occurring during these periods can affect early and/or late onset sepsis in low-resource African settings. Maternal and neonatal factors are represented separately with consideration of the health system factors and social, cultural and geo-political context known to influence bacterial infections. This framework guided our choice of variables for the logistic regression models.

**Figure 2.**
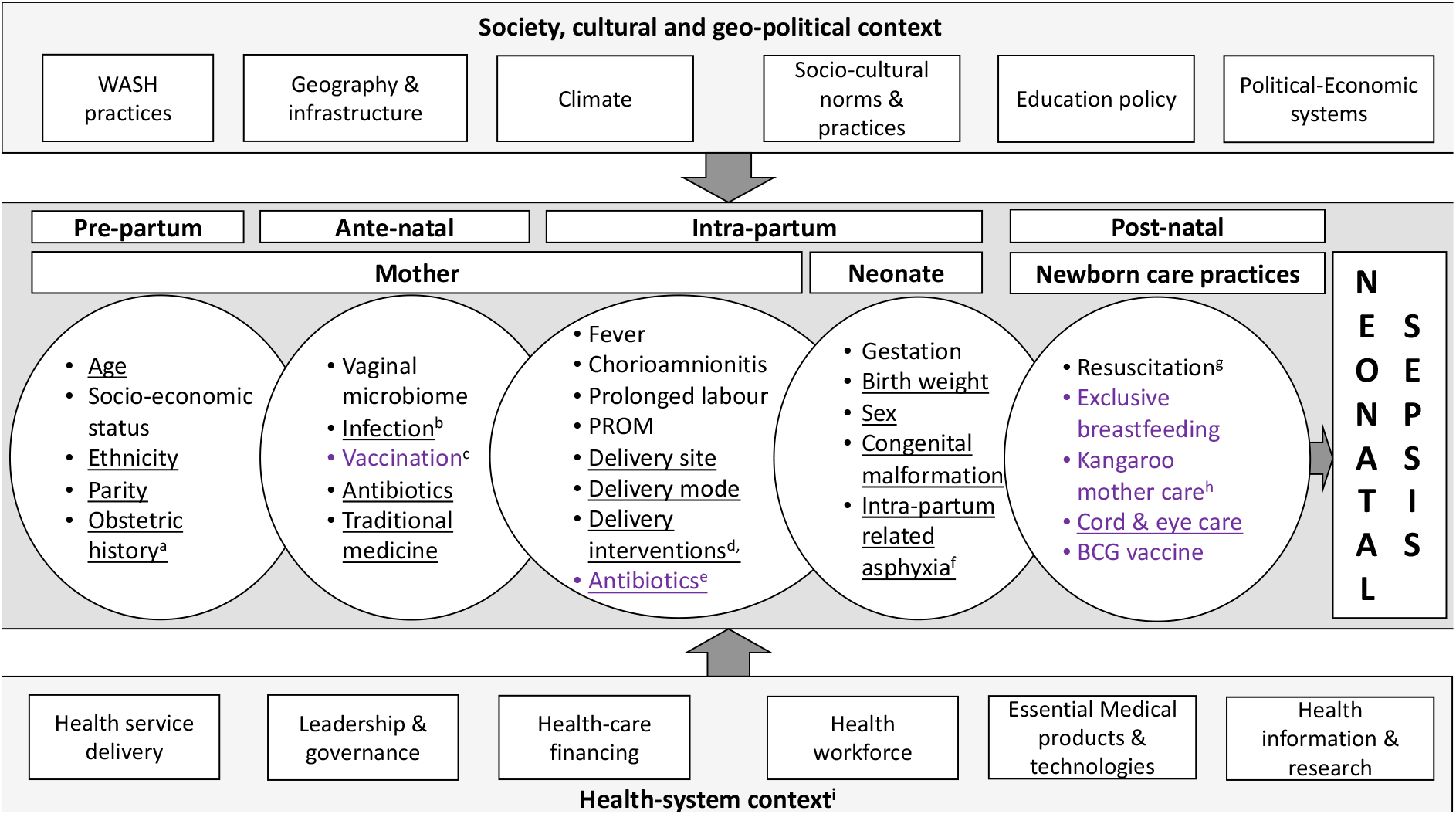
Conceptual framework to understand factors influencing the development of neonatal sepsis in low-resource West African settings. Variables which are underlined were available for the analysis Purple = potentially protective factors for neonatal sepsis; Black = potentially harmful factors for neonatal sepsis a) Obstetric history refers to history of previous miscarriage and/or stillbirth; b) Maternal urinary tract infection, sepsis, pneumonia; c) Maternal tetanus toxoid vaccine; d) Delivery interventions include multiple digital vaginal examinations, artificial rupture of membranes, episiotomy, forceps, vacuum extraction, or management of obstetric complications such as vaginal tear or uterine rupture; e) Intrapartum antibiotic treatment of maternal fever, suspected sepsis or chorioamnionitis; f) Intrapartum related asphyxia as detected by a 1 or 5-minute apgar score <7 or meconium stained amniotic fluid; g) Newborn resuscitation at delivery includes oxygen, resuscitation with face masks, suction; h) Kangaroo mother care applies to low birth weight newborns only; i) Health system context is as per WHO health systems building blocks framework [32].

Logistic regression analysis was used to identify factors associated with neonatal sepsis by comparing neonates meeting criteria for sepsis against those not admitted to a health facility during the neonatal period (28 days). In the single event of a neonate being admitted multiple times, only the first admission was considered in all analyses, to avoid inclusion of partially treated sepsis. Univariate (unadjusted) logistic regression was conducted to initially assess the relationship between independent variables and neonatal sepsis. An adjusted logistic regression model was then developed for each independent co-variate with *p*<0.2 on unadjusted regression, with this threshold chosen to avoid unnecessarily restrictive parameters for the adjusted models.[16] In-order to avoid over adjusting for multiple variables on the same causal pathway [17], each model only included variables occurring before or at the same timepoint as the independent variable of interest (Figure 2). Statistical significance for the adjusted analyses was pre-determined at *p*<0.05. Missing data was <1% except for outcome data (3.8%) and a complete case analysis approach was used.

## Results

### Participant overview

Of 6,773 neonates born to 6,684 pregnant women recruited to the PregnAnZI-2 trial in The Gambia, 6515 neonates were included in this study (Figure 3). Included neonates were predominantly normal birth weight (median 3.1kg, interquartile range (IQR) 2.8-3.4), singletons (6354/6515, 97.5%), delivered vaginally (6341/6515, 97.3%) by midwives (5611/6515, 86.1%). Mothers were aged median 27 years (IQR 23 – 31) and one-quarter (24.6%, 1605/6515) were primiparous (Table 1).

**Table 1.**
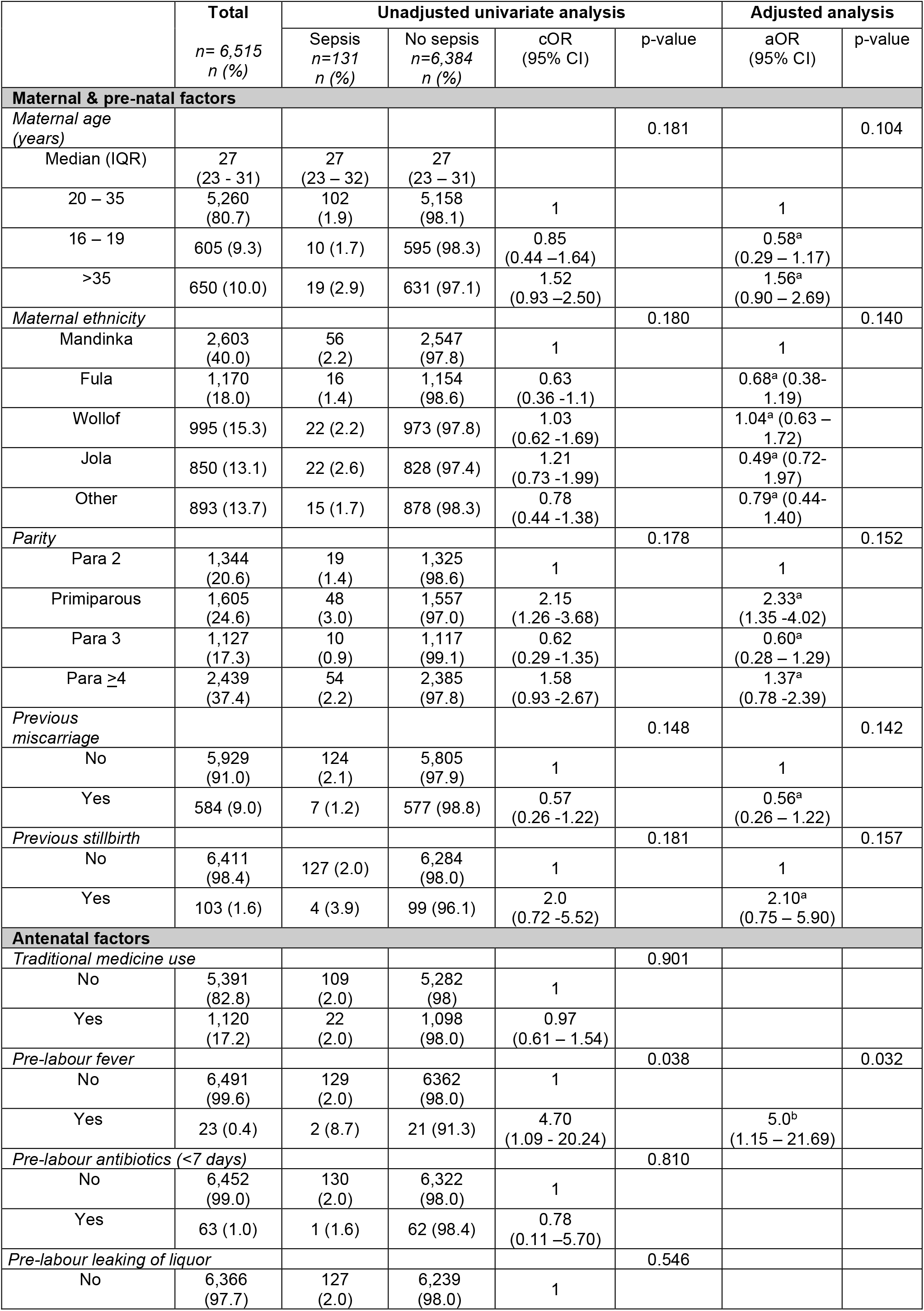
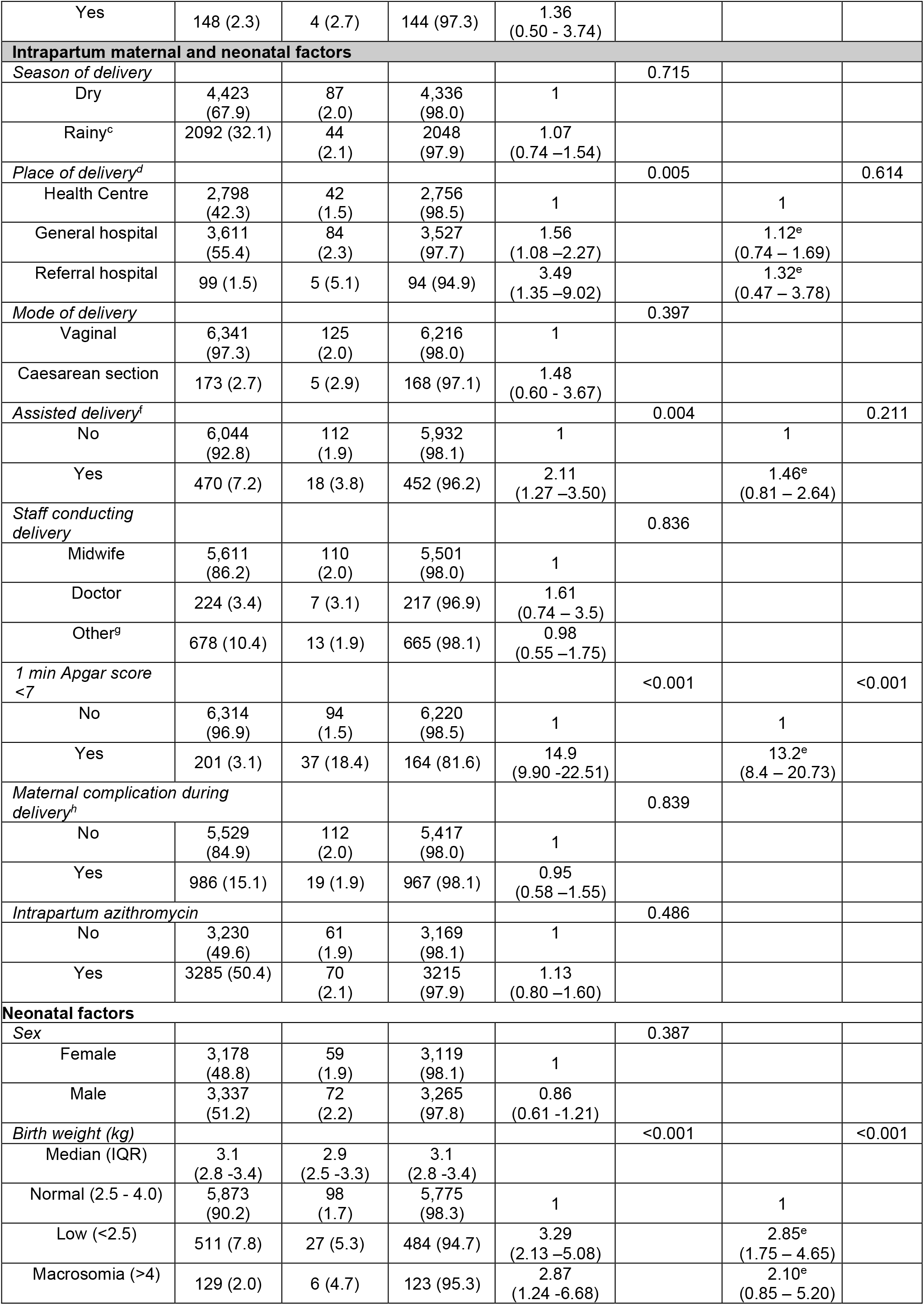
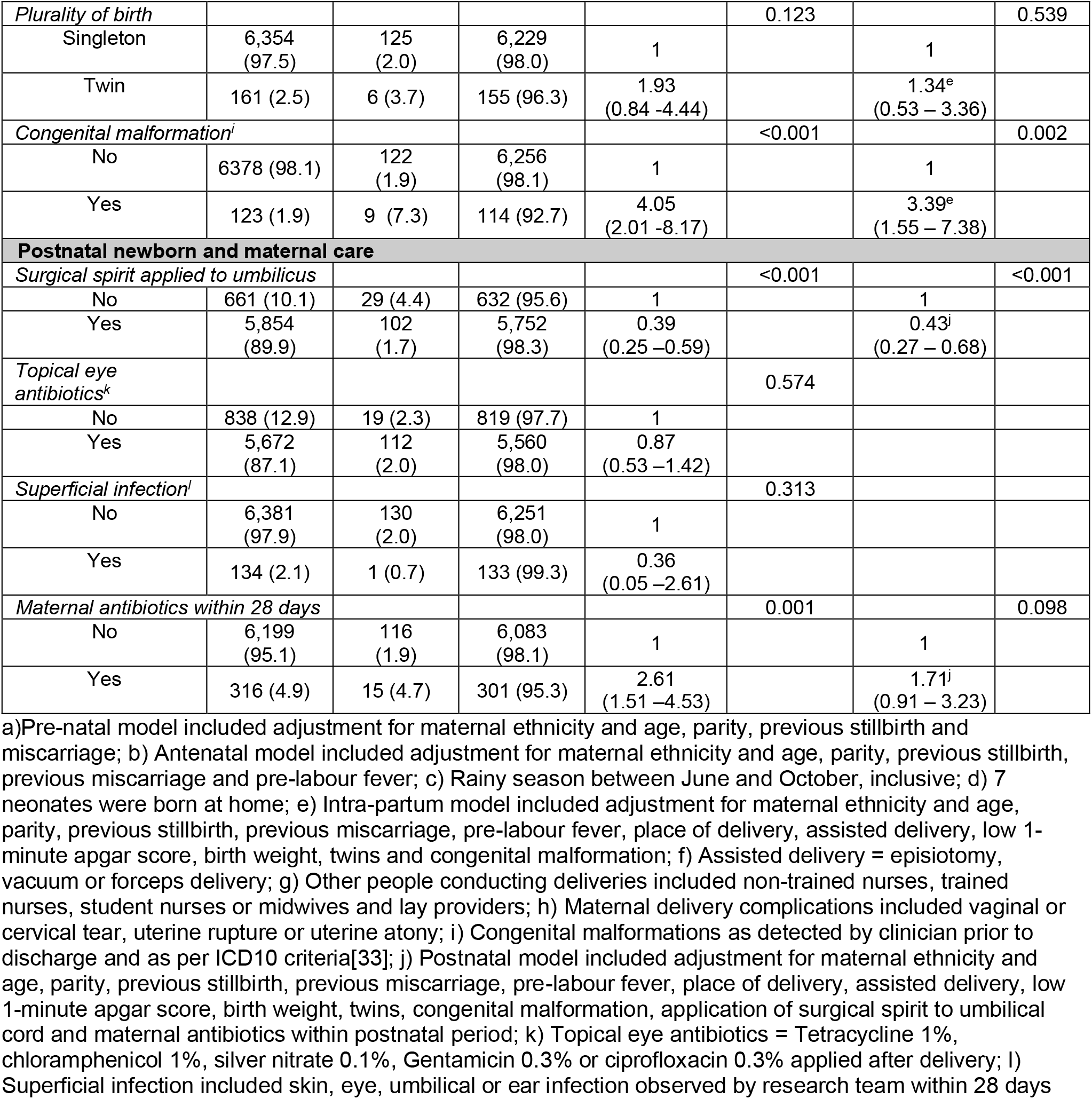
Maternal and neonatal sociodemographic and clinical factors associated with neonatal sepsis (suspected and confirmed cases)

**Figure 3.**
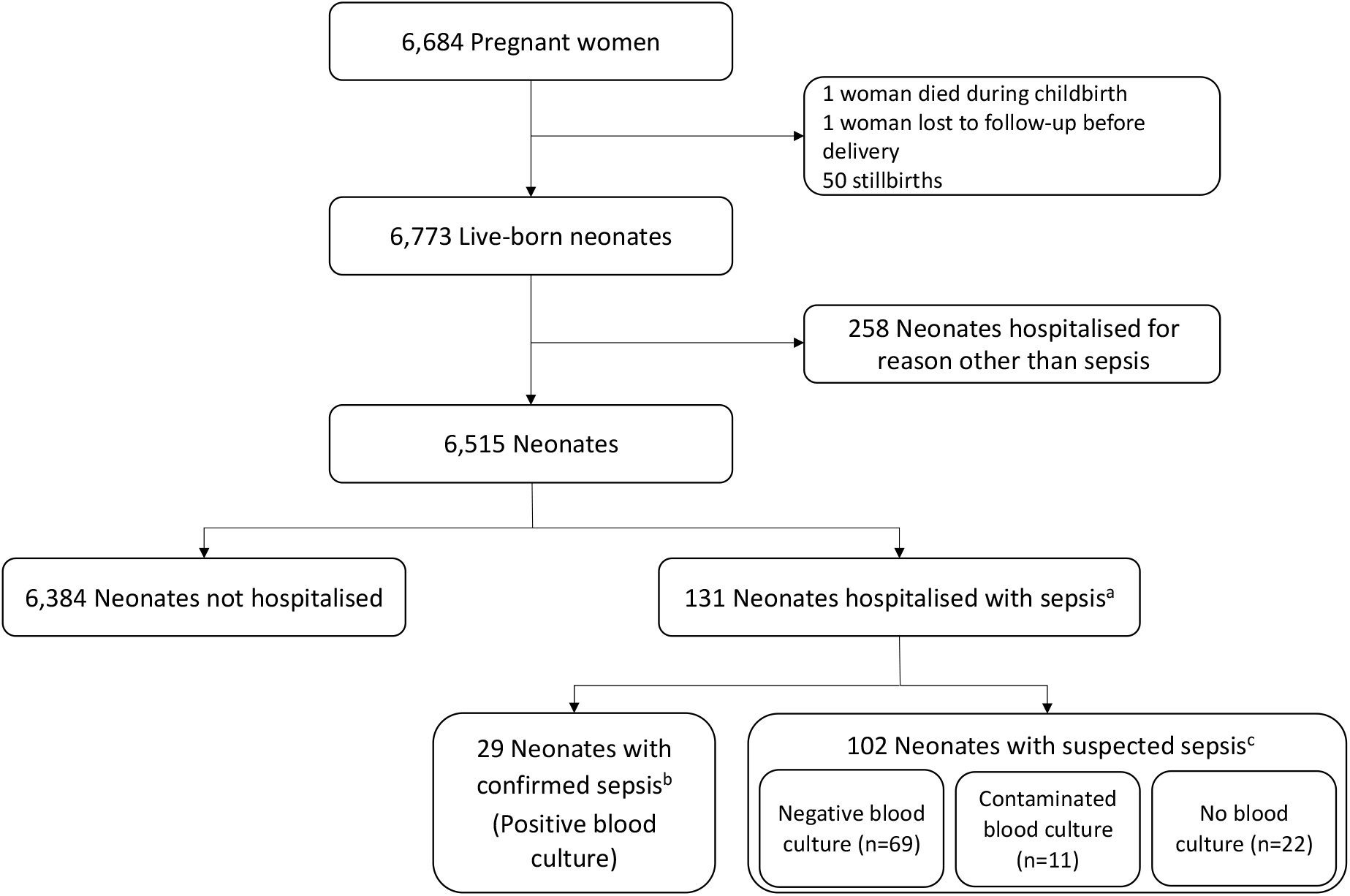
Flow chart indicating participant selection and including blood culture status for neonates with suspected sepsis. a) 1 neonate was admitted twice: Once with confirmed sepsis and subsequently with suspected sepsis; b) Blood cultures obtained due to clinical suspicion of sepsis by research physician, with 76% (22/29) meeting study diagnostic criteria for suspected sepsis; c) According to PregnAnZI-2 trial protocol criteria for sepsis (Figure 2)[11]

### Incidence of neonatal sepsis

131 neonates met criteria for sepsis (either suspected or confirmed sepsis), representing an overall incidence of 20.2 cases/1000 livebirths (131/6515). The incidence of early-onset sepsis was 15.7 cases/1000 livebirths (102/6515) compared to 4.5 cases/1000 livebirths (29/6515) for late-onset sepsis. Culture confirmed sepsis occurred in 22% (29/131) of cases (Figure 3), with an incidence of 4.5 cases/1000 livebirths (29/6515) for all confirmed sepsis and 2.8 cases/1000 livebirths for early-onset confirmed sepsis.

### Clinical features and inflammatory markers of sepsis case

Neonates with sepsis presented predominantly with respiratory distress (98/121, 81%), temperature instability (77/131, 59%) and gastrointestinal symptoms (59/122, 48%). 80% (105/131) of neonates had CRP level checked, of whom nearly two-thirds (63%, 66/105) had a raised CRP. A clinical suspicion of concomitant IRA, indicated by neonatal encephalopathy (NE) or meconium aspiration syndrome (MAS), was present in 29% (38/131) of neonates with sepsis. Neonates meeting criteria for sepsis but without a blood culture performed were more likely to have low 1-minute Apgar score (*p*=0.004), neurological symptoms (*p*=0.045), clinical suspicion of IRA as a co-morbidity (*p*<0.001)) and to die during the neonatal period (*p*=0.025) compared to neonates with confirmed or suspected sepsis who had a blood culture taken (Additional file 1).

### Aetiology of confirmed sepsis

11 bacterial species were identified from 29 neonates with confirmed sepsis, with no mixed growth. Gram-negative bacteria (18/29, 62%), were more commonly implicated than Gram-positive bacteria (11/29, 38%) with predominance during both early and late onset confirmed sepsis. *Burkholderia cepacia* (7/29, 24%) was the single most identified Gram-negative bacteria (Additional file 2). *Staph*y*lococcus aureus* was also responsible for 24% (7/29) of all confirmed sepsis and was the most common pathogen causing late-onset confirmed sepsis (45%, 5/11) (Additional file 2).

### Risk factors for neonatal sepsis

The only maternal factor associated with increased odds of neonatal sepsis was fever before onset of labour (aOR 5.00, 95% CI 1.15-21.69, *p*=0.032). Neonatal risk factors included IRA (measured as a low 1-minute Apgar score)(aOR 13.20, 95% CI 8.40 – 20.73, *p* <0.001), presence of an easily recognisable congenital malformation (aOR 3.39, 95% CI 1.55 – 7.38, *p*=0.002) and low birth weight (LBW) (aOR 2.85, 95% CI 1.75 - 4.65, *p* <0.001)(Table 1). Application of surgical spirit to the umbilical cord was associated with 57% reduced odds of sepsis (aOR 0.43, 95% 0.27-0.68, *p*<0.001) (Table 1). A sensitivity analysis excluding neonates who met criteria for suspected sepsis but lacked a blood culture showed no difference in variables associated with sepsis on the adjusted analysis (Additional file 3).

### Contribution of sepsis to neonatal mortality

40.7% (22/54) of all neonatal deaths in the cohort occurred in neonates with sepsis (Table 2) with a case-fatality rate (CFR) of 17% (22/127) for all (suspected and confirmed) sepsis (Additional file 1). Neonates with sepsis had a 40-fold increased odds of mortality during the first 28 days after delivery compared to neonates without sepsis (OR: 39.98, 95% CI 22.5 – 71.1, *p*<0.001)(Table 2).

**Table 2.**
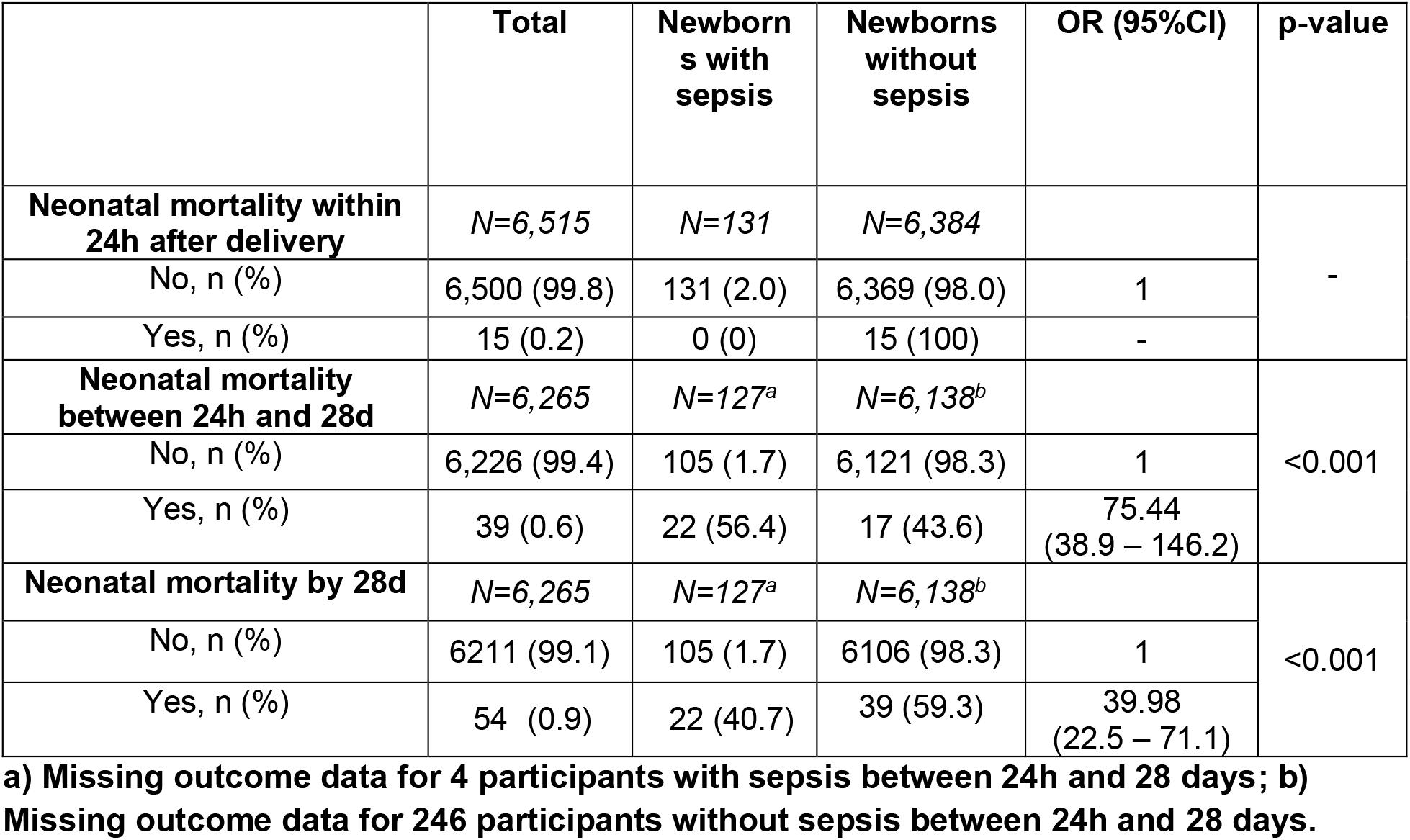
All-cause mortality amongst Gambian newborns with suspected or confirmed sepsis compared to neonates without sepsis.

|  | Total | Newborns with sepsis | Newborns without sepsis | OR (95%CI) | p-value |
| --- | --- | --- | --- | --- | --- |
| **Neonatal mortality within 24h after delivery** | *N*=6,515 | *N*=131 | *N*=6,384 |  | - |
| No, n (%) | 6,500 (99.8) | 131 (2.0) | 6,369 (98.0) | 1 |  |
| Yes, n (%) | 15 (0.2) | 0 (0) | 15 (100) | - |  |
| **Neonatal mortality between 24h and 28d** | *N*=6,265 | *N*=127<sup>a</sup> | *N*=6,138<sup>b</sup> |  | <0.001 |
| No, n (%) | 6,226 (99.4) | 105 (1.7) | 6,121 (98.3) | 1 |  |
| Yes, n (%) | 39 (0.6) | 22 (56.4) | 17 (43.6) | 75.44<br>(38.9 – 146.2) |  |
| **Neonatal mortality by 28d** | *N*=6,265 | *N*=127<sup>a</sup> | *N*=6,138<sup>b</sup> |  | <0.001 |
| No, n (%) | 6211 (99.1) | 105 (1.7) | 6106 (98.3) | 1 |  |
| Yes, n (%) | 54 (0.9) | 22 (40.7) | 39 (59.3) | 39.98<br>(22.5 – 71.1) |  |

a) Missing outcome data for 4 participants with sepsis between 24h and 28 days; b) Missing outcome data for 246 participants without sepsis between 24h and 28 days.

## Discussion

These secondary analyses of PregnAnZI-2 trial data identified a high incidence of sepsis, predominantly early onset, in 6515 neonates born following relatively healthy pregnancy and health facility delivery in The Gambia, West Africa. Although our cohort was not representative of all facility births, the use of an objective sepsis definition including both clinical and laboratory criteria, and the prospective collection of detailed data and comprehensive follow-up, enabled an in-depth analysis of risk factors guided by a context-specific conceptual framework. Risk factors for subsequent neonatal sepsis included fever prior to onset of labour, IRA (measured as 1-minute Apgar score <7), LBW and presence of an easily recognisable congenital malformation. Sepsis contributed substantially to neonatal mortality in our cohort, especially deaths occurring beyond the first 24h after delivery.

Neonatal sepsis incidence varies across Africa depending on geographical location, hospital versus community cohorts and population differences such as prevalence of LBW or maternal features such as HIV infection, which may alter the frequency of septic risk factors [18]. The health facility based incidence (20.2/1000 live births) of clinical sepsis in our cohort is high yet comparable to a large meta-analysis of Nigerian studies which collectively reported an incidence of 18.2/1000 live births (range 7-55) in >31,000 facility births from 9 observational studies conducted in urban secondary or tertiary level hospitals [19]. Comparison of our overall sepsis incidence with HIC settings is challenging due to differences in diagnostic criteria and thresholds for commencing antibiotics. However, our observed incidence of early onset confirmed sepsis (2.8/1000 livebirths) is over 5-fold higher than a large (n=757,979) cross-sectional hospital-based network study from Europe, North America and Australia (0.49/1000 live births) which utilised routine clinical and microbiological data [20]. With 15.48 million births reported in West Africa in 2022[21], our observed rates reflect an estimated neonatal sepsis burden of at least 312,000 infants annually in the region, with a conservative estimate of 5,300 neonatal deaths. Hence, addressing neonatal sepsis should be prioritised within child health policy in The Gambia and elsewhere in West Africa, with efforts focused on early onset sepsis prevention. This will require co-ordinated multi-disciplinary efforts across obstetric, neonatal and microbiology services with joined up policy and programmes to identify at risk women and newborns. Reducing neonatal sepsis and associated mortality is urgently needed for The Gambia to meet SDG 3.2 target by 2030, with health system strengthening required for improved sepsis diagnostics alongside high quality small and sick newborn care.

IRA was associated with 13-fold increased odds of neonatal sepsis in our cohort. This association between in-utero compromise and neonatal sepsis has previously been described in African newborns. A systematic review and meta-analysis of data from all African regions identified an association between several markers of IRA (meconium stained amniotic fluid, low 1- and 5-minute Apgar scores, resuscitation at birth and absent crying at birth) and neonatal sepsis, although there was high heterogeneity between studies[9]. One explanation is that perinatal infection may predispose to IRA[22] with emerging evidence of a complex inter-relationship between inflammation induced changes to the fetal immune system and perinatal brain injury [23]. This is supported by a detailed molecular and histological Ugandan case-control study demonstrating 8-fold increased odds of NE in the presence of early neonatal bacteraemia [24]. If this is the predominant pathway then maternal vaccination against specific pathogens causing sepsis (e.g., *E coli* or *Klebsiella pneumoniae*) or intra-partum antimicrobial strategies may have an impact on both neonatal sepsis and IRA in Africa. Alternatively, our finding may reflect early-onset sepsis due to horizontal transmission of pathogens from contaminated resuscitation equipment in the context of abundant environmental Gram-negative pathogens [25] and limited infection prevention control (IPC) strategies[26]. Mitigation of this pathway would require strengthening of facility IPC interventions and enhanced surveillance for nosocomial pathogens and clinical infection. A third possibility is over-estimation of sepsis amongst infants exposed to IRA, due to similarities in clinical presentation and similar rise in inflammatory markers in both common conditions. This is suggested by the different phenotype of neonates in our cohort who met sepsis criteria but who did not have a blood culture taken, with more neurological features, younger age, higher proportion with low 1-minute Apgar score and more clinical suspicion of NE or MAS, which are all consistent with IRA. Although there are many reasons for not obtaining a blood sample and we cannot verify why some neonates did not have blood taken for culture, it is possible that this group of neonates were severely unwell due to concomitant or isolated NE and obtaining large volumes of blood was not feasible. Despite the possibility that we overestimated sepsis cases, our sensitivity analysis excluding neonates without a blood culture identified the same risk factors on adjusted analysis. Further targeted research is required into the relationship between IRA and sepsis in different contexts, especially in low resource West African health facilities, in-order to inform prioritisation of intrapartum and immediate postnatal interventions and gain more insights to relative occurrence and interactions between these two important conditions. From our findings we also recommend that antibiotics should be provided as part of supportive care for Gambian neonates with NE due to IRA.

Our finding of an infection prevention effect from applying surgical spirit to the umbilical cord should be interpreted with caution as it may reflect reverse causality due to neonatal deaths occurring before hygienic cord care could be performed.

Gram-negative bacteria were the leading bacterial aetiology of neonatal sepsis in our cohort, consistent with a recent systematic review and meta-analysis reporting that 54% of blood-culture confirmed sepsis cases in West Africa are due to Gram-negative pathogens[7]. However, the contribution of Gram-negative versus Gram-positive pathogens in our cohort is not clear-cut, as the PregnAnZI-2 trial intervention (azithromycin) had proportionally more Gram-negative sepsis cases compared to the placebo arm, in which Gram-positive (predominantly *Staphylococcus aureus*) sepsis and Gram-negative sepsis were equivalent. Thus, we cannot exclude the possibility that azithromycin altered the pattern of sepsis aetiology by reducing *Staphylococcus aureus* sepsis.

*Burkholderia cepacia* was the most common Gram-negative bacteria identified in our cohort, with only two cases of *Klebsiella pneumoniae* sepsis. This contrasts with data from the multi-centre genomic surveillance study BARNARDS, which included an urban Nigerian site where *K pneumoniae* caused 6% of invasive (mostly late-onset) sepsis and no *Burkholderia s*pecies were detected [27]. A meta-analysis of neonatal sepsis aetiology also identified *Klebsiella* species as the most common Gram-negative causing bacteraemia or sepsis in West Africa, with *Burkholderia cepacia* not a dominant species[7]. However, in The Gambia, *Burkholderia cepacia* was recently implicated in endemic outbreaks affecting the national neonatal referral unit at EFSTH [25] and was a major cause of confirmed sepsis in term neonates born at either EFSTH or other regional hospitals with linkage to contaminated intravenous antibiotic and fluid preparations [28]. In our cohort, *Burkholderia cepacia* sepsis occurred mostly in neonates born and managed at a different health facility (BMCHH). Considered together, this evidence suggests that gram-negative pathogens, especially *Burkholderia cepacia*, are a common cause of neonatal sepsis in neonates born at urban western Gambian health facilities, possibly due to endemic or epidemic nosocomial outbreaks. IPC policy and practices at study sites should also be strengthened with further research needed to understand transmission pathways, especially between and within health facilities. Prospective neonatal sepsis surveillance is urgently needed at the trial sites, with a clinical-epidemiological outbreak investigation focused on *Burkholderia cepacia*. Our finding that *Staphylococcus aureus* most commonly causes late onset sepsis is consistent with other Gambian [28, 29] and West African studies [7, 27, 30], including a hospital-based case-control study of term neonates in the same Gambian region which reported *Staphylococcus aureus* as the predominant aetiology in neonates aged ≥7d with clinical sepsis [28]. The absence of Group B Streptococcus (GBS) sepsis in our cohort is also in keeping with previous West African studies [7, 19] and contrasts with Eastern and Southern Africa, where GBS is more commonly implicated, especially in early onset infections [7, 31]. Our insights into bacterial aetiology of neonatal sepsis supports the need for targeted local and national antimicrobial policies in The Gambia. Such policies should account for high risk of early onset sepsis due to Gram-negative pathogens with appropriate anti-staphylococcal antibiotic regimens for late-onset sepsis, incorporating local antimicrobial susceptibility data where possible.[29] We recognise that insights into bacterial aetiology may not be generalisable to other West African settings where *Burkholderia cepacia* is less prevalent or where the health system context or health-facility IPC provision differs. Our data indicates that sepsis is an important contributor to neonatal mortality with a case fatality rate (CFR) of 17% for all sepsis cases and 40-fold increased odds of mortality. Our reported CFR is lower than a similar Gambian hospital based neonatal study of possible serious bacterial infection which reported 30% CFR [28], although our study population had a lower proportion of LBW neonates, caesarean section deliveries and known acute or chronic maternal conditions, which may have altered the co-morbidity profile and neonatal mortality risk. A systematic review of neonatal sepsis in Nigerian hospitals reported an average overall mortality rate of 21.8% (12 studies conducted over past two decades), albeit with wide range (8% – 33%), likely reflecting heterogeneity in health system, population characteristics and temporal changes [19]. The “healthy effect” of participating in a clinical trial may have also influenced survival via enhanced surveillance for illness, additional education of families regarding danger signs and prompt clinical management, hence the mortality impact for non-research neonatal populations may be underestimated by our study.

Limitations of this post-hoc analysis include the exclusion of women with intra-partum fever, which is a known risk factor for neonatal sepsis [31]. Thus, estimates for sepsis incidence are conservative and relate only to the newborns of relatively healthy women without intrapartum septic risk factors. Conversely, despite using a sepsis definition which included both clinical and laboratory parameters, sepsis cases may have been over-estimated due to low specificity of criterion used and lack of diagnostics to exclude alternative or co-existing conditions such as viral respiratory infections or NE. This is a common challenge for neonatal sepsis research and more specific diagnostics are urgently needed, including host biomarkers capable of distinguishing between bacterial sepsis, viral infections and NE. Our findings relate to an urban population of health facility born neonates, but do not reflect the incidence, aetiology or risk factors for community born neonates with sepsis, which may differ.

## Conclusion

Sepsis, especially early-onset, is a major problem for hospital born neonates in urban Gambia with high associated mortality. We identified neonatal phenotypes (low birth weight, newborns with low apgar scores or those with easily recognisable congenital malformations) who may benefit from enhanced postnatal surveillance and targeted empirical antibiotic regimens covering Gram-negative (early onset) and *Staphylococcus aureus* (late onset) pathogens in-order to reduce avoidable neonatal mortality.

## Data Availability

Data are not publicly available. Qualified researchers may request access with the Gambia Government/MRCG Joint Ethics Committee. The review process and release of data will be facilitated by MRCG (http://www.mrc.gm/) through the Head of Governance. Access will not be unduly restricted.

## Abbreviations

aOR: Adjusted odds ratio
BCG: Bacillus Calmette-Guerin vaccine
BMCHH: Bundung Maternal and Child Health Hospital
CFR: Case fatality rate
CI: Confidence interval
CRP: C Reactive Protein
EDTA: Ethylene Diamine Tetra-acetic Acid
FBC: Full blood count
GBS: Group B Streptococcus
GCLP: Good Clinical Laboratory Practice
GNB: Gram-negative bacteria
HIC: High-income countries
HIV: Human Immunodeficiency Virus
IPC: Infection prevention control
IQR: Interquartile range
IRA: Intrapartum related asphyxia
LBW: Low birth weight
LMIC: Low- and middle-income countries
MAS: Meconium aspiration syndrome
MRCG: MRC Unit The Gambia at London School of Hygiene & Tropical Medicine
NE: Neonatal encephalopathy
OR: Odds ratio
ROC: Receiver Operating Characteristic
SDG: Sustainable Development Goal
WHO: World Health Organization

## Declarations

### Ethics approval and consent to participate

The PregnAnZI-2 trial was approved by The Gambia Government/Medical Research Council Unit The Gambia (MRCG) Joint Ethics Committee, the Comité d’Ethique pour la Recherche en Santé (CERS) and the Ministry of Health of Burkina Faso, and the LSHTM Ethics Committee. All women provided written informed consent for trial participation during antenatal care visits and and were free to withdraw at any time.

### Consent to publish

Not applicable.

### Authors’ contributions

AR, HT and UdA conceived and designed the PregnAnZI-2 trial. AR conceptualised this study with input from HB, CB, HT and UDA. Clinical data collection was carried out by UNN, NB, CJJ, FS, SG and MD with site co-ordination by BC. KM gave oversight to data collection at BMCHH site. Laboratory data was generated by the laboratory team (SD, SU, AB, ES, IJ, BC, MB, KK, AC). YN and EN oversaw database design, cleaning and preparation for analysis. HB performed the analysis with full access to the data and input from AR. HB drafted the initial manuscript with input to intellectual content from AR, UDA, HT, CB, UN, SG, NB and BC. All authors have read and approved the final version. AR gave oversight to the work as guarantor and accepts full responsibility for finished work and controlled the decision to publish.

## Acknowledgements

We acknowledge the support of the Gambian Government Ministry of Health and all health facility management and health personnel who were involved at the study sites (Serekunda Health Centre and Bundung Maternal and Child Health Hospital) and referral sites (Edward Francis Small Teaching Hospital and Kanifing General Hospital). We acknowledge the inputs from Clinical Services Department at MRC Gambia, along with all research support provided by MRCG. Thank you to the patients and their families who took part in this study.

