## Supplementary Table 1 for "Incidence, risk factors and outcomes for neonatal sepsis in The Gambia: Descriptive cohort analysis embedded in a randomised clinical trial"

Supplementary Table 1. Clinical and laboratory features of neonatal sepsis stratified by blood culture status from a cohort of Gambian neonates

| **Characteristic** | **Total sepsis**  **(N=131)** | **Confirmed sepsis**  **Positive blood culture**  **(N=29)** | **Suspected sepsis (N=102)** | | | ***P* value^a^** |
| --- | --- | --- | --- | --- | --- | --- |
|  |  |  | **Negative blood culture**  **(N=69)** | **Contaminated blood culture**  **(N=11)** | **No blood culture**  **(N=22)** |  |
| Male sex, N^o^ (%) | 72 (55) | 16 (55) | 39 (57) | 3 (27) | 14 (64) | 0.482 |
| Admission age, h (median, IQR) | 19 (5-50) | 35 (9 – 177) | 20 (8 – 32) | 23 (17 – 128) | 4 (2 – 7) | 0.670 |
| Age <72h at admission, N^o^ (%) | 102/129 (79) | 18 (62) | 58/68 (85) | 8 (73) | 18/21 (86) | 0.563 |
| Birth weight (median, IQR) | 2.9 (2.5 – 3.3) | 2.8 (2.5 – 3.1) | 2.9 (2.6 – 3.3) | 3.3 (2.9 – 3.4) | 2.9 (2.3 – 3.2) | 0.744 |
| Low-1 min apgar score, *N^o^ (%)* | 37 (28) | 5 (17) | 18 (26) | 2 (18) | 12 (55) | 0.004 |
| **Clinical signs of sepsis** | | | | | | |
| Respiratory distress,^b^ *N^o^ (%)* | 98/121 (81) | 20/27 (74) | 53/64 (83) | 7/10 (70) | 18/20 (90) | 0.359 |
| Fever or hypothermia,^c^ *N^o^ (%)* | 77 (59) | 18 (62) | 40 (58) | 5 (45) | 14 (64) | 0.644 |
| Lethargy/irritability, *N^o^ (%)* | 29 (22) | 7 (24) | 14 (20) | 3 (27) | 5 (23) | 1.0 |
| Gastrointestinal symptoms,^d^ *N^o^ (%)* | 59/122 (48) | 14/28 (50) | 34/65 (52) | 8/10 (80) | 3/19 (16) | 0.002 |
| Bleeding, *N^o^ (%)* | 2/122 (2) | 1/28 (4) | 1/65 (2) | 0 | 0 | 1.0 |
| Central nervous system symptoms,^e^ *N^o^ (%)* | 22/121 (18) | 3/28 (11) | 10/65 (15) | 2/9 (22) | 7/19 (37) | 0.045 |
| **Laboratory signs of sepsis** | | | | | | |
| Elevated C-reactive protein,^f^ *N^o^ (%)* | 66/105^g^ (63) | 11/21 (52) | 40/61 (66) | 6/7 (86) | 9/16 (56) | 0.583 |
| Abnormal white blood cell count,^h^ *N^o^ (%)* | 34/123 (28) | 6/28 (21) | 17/64 (27) | 3/10 (30) | 8/20 (40) | 0.183 |
| Abnormal neutrophil count,^i^ *N^o^ (%)* | 26/113 (23) | 6/25 (24) | 11/59 (19) | 2/10 (20) | 7/19 (37) | 0.138 |
| Low platelet count,^j^ *N^o^ (%)* | 35/122 (29) | 5/28 (18) | 20/64 (31) | 5/10 (50) | 5/20 (25) | 0.792 |
| **Co-morbidities and outcome** | | | | | | |
| Clinical suspicion of intra-partum related asphyxia,^k^ *N^o^ (%)* | 38 (29) | 4 (14) | 19 (28) | 1/10 (10) | 14 (64) | <0.001 |
| Case fatality rate,^l^ *N^o^ (%)* | 22/127 (17) | 5 (17) | 8^l^ (12) | 1^l^ (10) | 8 (36) | 0.025 |

a) Statistical difference between neonates with blood culture taken (n=109) versus no blood culture taken (n=22); b) Respiratory rate >60 breaths/min, cyanosis, chest wall indrawing, grunting, respiratory distress noted in medical records; c) Axillary temperature >38.0^o^C (not due to external warming), or axillary temperature <36.0^o^C; d) Abdominal distension, feeding intolerance (>20% residual aspirate over 24h), poor feeding after feeding well, ≥2 episodes of vomiting; e) Seizures, bulging fontanelle, single witnessed episode of apnoea; f) CRP >10 mg/l (Early onset sepsis) or >40 mg/l (late onset sepsis); g) Missing data on CRP levels for 26 neonates due to insufficient blood volumes (n=17) and protocol deviations in obtaining CRP levels for late-onset sepsis (n=9); h) WBC <5 x10^9^/l or >25 x10^9^/l in absence of receiving corticosteroids; i) Absolute neutrophil count <1.75 x10^9^/l or >15 x10^9^/l; j) Platelet count <150 x10^9^/l; k) Either neonatal encephalopathy or meconium aspiration syndrome, as diagnosed by research clinician using clinical discretion; l) Outcome missing for 4 newborns, of which 3 had a negative blood culture and 1 had a contaminated culture
