## Supplementary Table 2 for "Incidence, risk factors and outcomes for neonatal sepsis in The Gambia: Descriptive cohort analysis embedded in a randomised clinical trial"

Supplementary Table 2. Bacterial aetiology of confirmed sepsis in Gambian hospital-born neonates

| Bacteria | Total sepsis  N=29  N (%) | Early onset sepsis (0-72h)  N=18  N (%) | Late onset sepsis (72h-28d)  N=11  N (%) |
| --- | --- | --- | --- |
| Gram Positive | | |  |
| *Staphylococcus aureus*^a^ | 7 (24) | 2 (11) | 5 (45) |
| *Corynebacterium propinquum* | 2 (7) | 2 (11) | 0 |
| Enterococcus species | 2 (7) | 2 (11) | 0 |
| TOTAL GRAM POSITIVE | 11 (38) | 6 (33) | 5 (45) |
| Gram Negative | | | |
| *Burkholderia cepacia* | 7 (24) | 6 (33) | 1 (9) |
| *Escherichia coli*^a^ | 3 (10) | 0 | 3 (27) |
| *Klebsiella pneumoniae* | 2 (7) | 1 (6) | 1 (3) |
| Enterobacter species^a,b^ | 2 (7) | 2 (11) | 0 |
| Pseudomonas species | 1 (3) | 1 (6) | 0 |
| *Citrobacter youngae* | 1 (3) | 1 (6) | 0 |
| *Hafnia alvei* | 1 (3) | 1 (6) | 0 |
| *Weeksella virosa* | 1 (3) | 0 | 1 (3) |
| TOTAL GRAM NEGATIVE | 18 (62) | 12 (67) | 6 (55) |

*a)* Five neonates who died had culture confirmed sepsis. Of these, 2 had *Staphylococcus aureus,* 2 *Enterobacter* spp. and 1 *Escherichia coli*.

*b) Enterobacter cloacae* (n=1) and *Enterobacter kobei* (n=1)
