## Supplementary Table 3 for "Incidence, risk factors and outcomes for neonatal sepsis in The Gambia: Descriptive cohort analysis embedded in a randomised clinical trial"

Supplementary Table 3. Sensitivity analysis to identify risk factors for neonatal sepsis, excluding neonates with no blood culture

|  | **Total**  *n= 6,493*  *n (%)* | **Unadjusted univariate analysis** | | | | **Adjusted analysis** | |
| --- | --- | --- | --- | --- | --- | --- | --- |
|  |  | Sepsis  *n=109*  *n (%)* | No sepsis  *n=6,384*  *n (%)* | cOR  (95% CI) | p-value | aOR  (95% CI) | p-value |
| **Maternal & pre-natal factors** | | | | | | | |
| *Maternal age (years)* |  |  |  |  | 0.349 |  |  |
| Median (IQR) | 27 (23-31) | 27(24-32) | 27 (23 – 31) |  |  |  |  |
| 20 – 35 | 5,244 (80.8) | 86 (1.6) | 5,158 (98.4) | 1 |  |  |  |
| 16 – 19 | 603 (9.3) | 8 (1.3) | 595 (98.7) | 0.81 (0.39-1.67) |  |  |  |
| >35 | 650 (10.0) | 19 (2.9) | 631 (97.7) | 1.43 (0.82-2.48) |  |  |  |
| *Maternal ethnicity* |  |  |  |  | 0.279 |  |  |
| Mandinka | 2,593 (40.0) | 46 (1.8) | 2,547 (98.2) | 1 |  |  |  |
| Fula | 1,166 (18.0) | 12 (1.0) | 1,154 (99.0) | 0.58 (0.30-1.09) |  |  |  |
| Wollof | 992 (15.3) | 19 (1.9) | 973 (98.1) | 1.08 (0.63-1.85) |  |  |  |
| Jola | 846 (13.0) | 18 (2.1) | 828 (97.9) | 1.20 (0.69-2.09) |  |  |  |
| Other | 892 (13.7) | 14 (1.6) | 878 (98.4) | 0.88 (0.30-1.09) |  |  |  |
| *Parity* |  |  |  |  | 0.310 |  |  |
| Para 2 | 1,339 (20.6) | 14 (1.0) | 1,325 (99.0) | 1 |  |  |  |
| Primiparous | 1,597 (24.6) | 40 (2.5) | 1,557 (97.5) | 2.43 (1.32-4.49) |  |  |  |
| Para 3 | 1,126 (17.3) | 9 (0.8) | 1,117 (99.2) | 0.76 (0.33-1.77) |  |  |  |
| Para ≥4 | 2,431 (37.4) | 46 (1.9) | 2,385 (98.1) | 1.83 (1.0-3.33) |  |  |  |
| *Previous miscarriage* |  |  |  |  | 0.206 |  |  |
| No | 5,908 (91.0) | 103 (1.7) | 5,805 (98.3) | 1 |  |  |  |
| Yes | 583 (9.0) | 6 (1.0) | 577 (99.0) | 0.59 (0.26-1.34) |  |  |  |
| *Previous stillbirth* |  |  |  |  | 0.089^a^ |  |  |
| No | 6,389 (98.4) | 105 (1.6) | 6,284 (98.4) | 1 |  |  |  |
| Yes | 103 (1.6) | 4 (3.9) | 99 (96.1) | 2.42 (0.87-6.69) |  |  |  |
| **Antenatal factors** | | | | | | | |
| *Traditional medicine use* | |  |  |  | 0.952 |  |  |
| No | 5,372 (82.8) | 90 (1.7) | 5,282 (98.3)) | 1 |  |  |  |
| Yes | 1,117 (17.2) | 19 (1.7) | 1,098 (98.3) | 1.02 (0.62-1.67) |  |  |  |
| *Pre-labour fever* |  |  |  |  | 0.020 |  | 0.019 |
| No | 6.469 (99.6) | 107 (1.7) | 6,362 (98.3) | 1 |  |  |  |
| Yes | 23 (0.4) | 2 (8.7) | 21 (91.3) | 5.66 (1.31-24.45) |  | 5.79^b^ (1.34 – 25.02) |  |
| *Pre-labour antibiotics (<7 days)* | |  |  |  | 0.955 |  |  |
| No | 6,430 (99.0) | 108 (1.7) | 6,322 (98.3) | 1 |  |  |  |
| Yes | 63 (1.0) | 1 (1.6) | 62 (98.4) | 0.944 (0.13-6.87) |  |  |  |
| *Pre-labour leaking of liquor* | |  |  |  | 0.730 |  |  |
| No | 6,345 (97.7) | 106 (1.7) | 6,239 (98.3)) | 1 |  |  |  |
| Yes | 147 (2.3) | 3 (2.0) | 144 (98) | 1.23 (0.38-3.91) |  |  |  |
| **Intrapartum maternal and neonatal factors** | | | | | | | |
| *Season of delivery* |  |  |  |  | 0.834 |  |  |
| Dry | 4,409 | 73 (1.7) | 4,336 (98.3) | 1 |  |  |  |
| Rainy^c^ | 2084 (32.1) | 36 (1.7) | 2048 (98.3) | 1.04 (0.70-1.56) |  |  |  |
| *Place of delivery^d^* |  |  |  |  | 0.048 |  | 0.807 |
| Health Centre | 2,792 (43.0) | 36 (1.3) | 2,756 (98.7) | 1 |  | 1 |  |
| General hospital | 3,598 (55.4) | 71 (2.0) | 3,527 (98.0) | 1.54 (1.03-2.3) |  | 1.17^e^ (0.76 – 1.79) |  |
| Referral hospital | 96 (1.5) | 2 (2.1) | 94 (97.9) | 1.63 (0.39-6.87) |  | 0.76^e^ (0.17 – 3.46) |  |
| *Mode of delivery* |  |  |  |  | 0.925 |  |  |
| Vaginal | 6,321 (97.4) | 105 (1.7) | 6,216 (98.3) | 1 |  |  |  |
| Caesarean section | 171 (2.6) | 3 (1.8) | 168 (98.2) | 1.06 (0.33-3.36) |  |  |  |
| *Assisted delivery*^f^ |  |  |  |  | 0.110 |  | 0.255 |
| No | 6,028 (92.9) | 96 (1.6) | 5,932 (98.4) | 1 |  | 1 |  |
| Yes | 464 (7.1) | 12 (2.6) | 452 (97.4) | 1.64 (0.89-3.01) |  | 1.44^e^ (0.77 – 2.71) |  |
| *Staff conducting delivery* |  |  |  |  | 0.900 |  |  |
| Midwife | 5593 (86.2) | 92(1.6) | 5,501 (98.4) | 1 |  |  |  |
| Doctor | 222 (3.4) | 5 (2.3) | 217 (97.7) | 1.38 (0.55-3.42) |  |  |  |
| Other^g^ | 676 (10.4) | 11 (1.6) | 665 (98.4) | 0.99 (0.53-1.86) |  |  |  |
| *1 min Apgar score <7* |  |  |  |  | <0.001 |  | <0.001 |
| No | 6,304 (97.1) | 84 (1.3) | 6,220 (98.7) | 1 |  | 1 |  |
| Yes | 189 (2.9) | 25 (13.2) | 164 (86.8) | 11.29 (7.04-18.11) |  | 10.26^e^ (6.18– 17.06) |  |
| *Maternal complication during delivery^h^* | |  |  |  | 0.892 |  |  |
| No | 5,510 (84.9) | 93 (1.7) | 5,417 (98.3) | 1 |  |  |  |
| Yes | 983 (15.1) | 16 (1.6) | 967 (98.4) | 0.96 (0.56-1.65) |  |  |  |
| *Intrapartum azithromycin* | |  |  |  | 0.555 |  |  |
| No | 3,220 (49.6) | 51 (1.6) | 3,169 (98.4) | 1 |  |  |  |
| Yes | 3273 (50.4) | 58 (1.8) | 3215 (98.2) | 1.12 (0.77-1.64) |  |  |  |
| **Neonatal factors** | | | | | | | |
| *Sex* |  |  |  |  | 0.669 |  |  |
| Female | 3,170 (48.8) | 51 (1.6) | 3,119 (98.4) | 1 |  |  |  |
| Male | 3,323 (51.2) | 58 (1.7) | 3,265 (98.3) | 0.92 (0.63-1.35) |  |  |  |
| *Birth weight (kg)* |  |  |  |  | <0.001 |  | 0.002 |
| Median (IQR) | 3.1 (2.8-3.4) | 2.9 (2.5-3.3) | 3.1 (2.8-3.4) |  |  |  |  |
| Normal (2.5 - 4.0) | 5,859 (90.3) | 84 (1.4) | 5,775 (98.6) | 1 |  | 1 |  |
| Low (<2.5) | 505 (7.8) | 21 (4.2) | 484 (95.8) | 2.98 (1.83-4.85) |  | 2.62^e^ (1.57 – 4.39) |  |
| Macrosomia (>4) | 127 (2.0) | 4 (3.1) | 123 (96.9) | 2.24 (0.81-6.19) |  | 1.75^e^ (0.61 – 5.06) |  |
| *Plurality of birth* |  |  |  |  | 0.409 |  |  |
| Singleton | 6,334 (97.6) | 105 (1.7) | 6,229 (98.3) | 1 |  |  |  |
| Twin | 159 (2.4) | 4 (2.5) | 155 (97.5) | 1.53 (0.56-4.21) |  |  |  |
| *Congenital malformation^i^* | |  |  |  | <0.001 |  | 0.001 |
| No | 6,357 (98.1) | 101 (1.6) | 6,256 (98.4) | 1 |  | 1 |  |
| Yes | 122 (1.9) | 8 (6.6) | 114 (93.4) | 4.35 (2.07-9.14) |  | 3.75^e^ (1.69 – 8.32) |  |
| **Postnatal newborn and maternal care** | | | | | | | |
| *Surgical spirit applied to umbilicus* | |  |  |  | <0.001 |  | 0.001 |
| No | 656 (10.1) | 24 (3.7) | 632 (96.3) | 1 |  | 1 |  |
| Yes | 5,837 (89.9) | 85 (1.5) | 5,752 (98.5) | 0.39 (0.25-0.62) |  | 0.45^j^ (0.28 – 0.74) |  |
| *Topical eye antibiotics^k^* |  |  |  |  | 0.778 |  |  |
| No | 832 (12.8) | 13 (1.6) | 819 (98.4) | 1 |  |  |  |
| Yes | 5,656 (87.2) | 96 (1.7) | 5,560 (98.3) | 1.09 (0.61-1.95) |  |  |  |
| *Superficial infection^l^* |  |  |  |  | 0.409 |  |  |
| No | 6,359 (97.9) | 108 (1.7) | 6,251 (98.3) | 1 |  |  |  |
| Yes | 134 (2.1) | 1 (0.7) | 133 (99.3) | 0.44 (0.06-3.14) |  |  |  |
| *Maternal antibiotics within 28 days* | |  |  |  | 0.003 |  | 0.049 |
| No | 6,199 (95.1) | 116 (1.9) | 6,083 (98.1) | 1 |  | 1 |  |
| Yes | 313 (4.8) | 15 (3.8) | 301 (96.2) | 2.50 (1.36-4.61) |  | 1.96^j^ (1.0 – 3.83) |  |

a) Previous stillbirth not adjusted, as no other maternal or prenatal variables significantly associated on unadjusted analysis ; b) Antenatal model included adjustment for parity, previous stillbirth and pre-labour fever; c) Rainy season between June and October, inclusive; d) 7 neonates were born at home; e) Intra-partum model included adjustment for parity, previous stillbirth, pre-labour fever, place of delivery, assisted delivery, low 1-minute apgar score, birth weight and congenital malformation; f) Assisted delivery = episiotomy, vacuum or forceps delivery; g) Other people conducting deliveries included auxillary nurses, trained nurses, student nurses or midwives and lay providers; h) Maternal delivery complications include vaginal or cervical tear, uterine rupture or uterine atony; i) Congenital malformations as detected by clinician prior to discharge and as per ICD10 criteria(35); j) Post-natal model included adjustment for parity, previous stillbirth, pre-labour fever, place of delivery, assisted delivery, low 1-minute apgar score, birth weight and congenital malformation, application of surgical spirit to umbilical cord and maternal antibiotics within postnatal period; k) Topical eye antibiotics = Tetracycline 1%, chloramphenicol 1%, silver nitrate 0.1%, Gentamicin 0.3% or ciprofloxacin 0.3% applied after delivery; l) Superficial infection included skin, eye, umbilical or ear infection observed by research team within 28 days
